# Identification of hippocampal volume as a mediator of the association between *APOE4* and dementia

**DOI:** 10.1101/2020.12.04.20242461

**Authors:** Sixtine Brenek, Stéphanie Debette, Sophie Auriacombe, Philippe Amouyel, Vincent Chouraki, Aline Meirhaeghe, Vincent Damotte

## Abstract

**Objective:** To identify any mediating effect of vascular and neurodegenerative risk factors of dementia in the association existing between *APOE4* and dementia.

**Methods:** 1,240 participants from the French Three City Dijon Study without prevalent tumor or dementia were included. Among these participants, 76 developed dementia during the 12 years of follow-up. Using regression and mediation analyses, we studied whether known risk factors for dementia i.e smoking status, body mass index, diabetes mellitus, hypercholesterolemia, hypertension, hippocampal volume, rate of hippocampal volume loss or white matter hyperintensities could mediate the association between *APOE4* and dementia. Regression models were adjusted for age, sex, education level (and total intracranial volume when imaging metrics were considered).

**Results:** Hippocampal volume was a partial mediator of the association between *APOE4* and dementia (mediation ratio = 6.7%, p=0.03). No mediation effect was found for hypercholesterolemia. No mediation analyses could be undertaken for the others factors due to the lack of their association with *APOE4* in our sample.

**Conclusions:** In this study, the association between *APOE4* and dementia was partially mediated by hippocampal volume, confirming a deleterious role of *APOE4* in the hippocampus. This result warrants further replication in other cohorts, with higher sample sizes.

## Introduction

Dementia is a clinical syndrome characterized by impairment of memory, language, and behavior. While Alzheimer’s disease (AD) is the most common subtype of dementia, several other types can occur such as vascular dementia, mixed dementia, dementia with Lewy bodies (LBD) and Parkinson’s disease (PD) dementia^1^.

The *Apolipoprotein E ε4* allele (*APOE4*) is a major genetic risk factor for AD^2, 3^. Studies have also shown an association between *APOE* genotype and the risk of LBD and PD dementia^3^. *APOE4* was also found to be associated with a lower hippocampal volume (HV) and a higher HV atrophy rate in AD patients^4, 5^, an increased risk of hypertension^6, 7^, hypercholesterolemia^8^ and a lower body mass index (BMI)^9^. A trend for higher white matter hyperintensities (WMH) burden in *APOE4* carriers has also been observed^10^.

In addition, several vascular risk factors can also influence the risk of dementia: age, hypercholesterolemia, hypertension, diabetes and smoking^11^. Neurodegenerative factors such as WMH^12^ and HV^13^ are also associated with risk of dementia. Protective factors have also been identified such as high education level^11^.

The importance of the role of specific vascular and neurodegenerative risk factors in cognition was emphasized recently. A mediation analysis showed that the association of microvascular lesions and neurodegeneration score with cognitive decline was mainly attributed to WMH changes, and to a much less extent, to gray matter volume^14^. In this context, we aimed at quantifying the mediating effect of several vascular and neurodegenerative risk factors on the association between *APOE4* and the risk of dementia.

## Materials and methods

### Population

Participants included in this study are from the Three-City (3C) study, an on-going population-based prospective cohort designed to examine the relationship between vascular diseases and dementia in 9,294 persons aged 65 years and older ^15^. Participants were recruited from 1999 to 2001 in three French cities: Bordeaux, Montpellier and Dijon. The ethics committee of the Kremlin-Bicêtre hospital approved the 3C protocol and all participants signed an informed consent. Only participants with available MRI data at baseline and at 4 years of follow-up, all recruited in Dijon, were included in this study (n=1,434). Participants with a brain tumor at baseline, prevalent dementia, missing *APOE4* genotype or any variables used as covariates in the analysis (age, sex, education level, total intracranial volume (TIV)) were excluded from analysis (n=194). Two independent analyses were performed, considering mediators assessed at baseline or mediators assessed at 4 years of follow-up. In the second analysis using the mediators assessed at 4 years of follow-up, participants who developed a dementia between the baseline visit and the visit at 4 years were removed (n=7).

### Outcome

The outcome considered was the incident dementia status after up to 12 years of follow-up. All types of dementia were considered. Diagnosis was based on the criteria of the Diagnostic and Statistical Manual of Mental Disorders (Fourth Edition) and is fully described elsewhere 16, 17.

### *APOE4* allele

Genotyping of *APOE4* was already performed in 3C Study participants and the protocol is fully described elsewhere ^18^. *APOE4* was coded as 0, 1 or 2 according to the number of *APOE4* alleles carried by individuals.

### Mediators

The following variables were studied as potential mediators of the association between *APOE4* and dementia: hypertension, WMH volume (in cm3) and HV (in cm3), assessed at baseline and at 4 years of follow-up as well as atrophy rate of HV (in cm3/year), computed as: atrophy rate = (HV_4years_ – HV_baseline_)/(t _4years_ – t _baseline)._ BMI (in kg/m^2^), hypercholesterolemia, diabetes and smoking status at baseline were also studied as mediators. Hypertension was defined as having a systolic blood pressure higher than 140 mmHg or a diastolic blood pressure lower than 90 mmHg, or by using hypertension treatment. Hypercholesterolemia was defined as having a total cholesterol level higher than 6.2 mmol/L or by using lipid-lowering medication. Diabetes was defined as having a glycemia higher than 7mmol/L or by using an antidiabetic medication. Smoking status was considered under three different models (current vs never smoker, current versus former/never smoker and current/former versus never smoker). All structural brain scans were acquired using the same MRI machine (1.5 T; Siemens, Erlangen) and the same protocol. The three-dimensional (3D) high-resolution T1-weighted brain volume and T2-weighted brain volume were acquired and processed as described elsewhere ^19-22^. WMH variable was log-transformed to account for its non-normal distribution.

Presence of outliers in BMI, TIV, WMH, HV and HV atrophy rate variables was assessed using the interquartile range method. Eventually, after removing missing values and/or outliers, analyses at baseline were performed on 1,240 participants for smoking status, hypercholesterolemia and hypertension, 1,238 participants for diabetes, 1,223 participants for BMI, 1,221 and 1,197 participants for HV and HV atrophy rate, respectively (by also excluding outliers for TIV) and 955 participants for WMH (by also excluding outliers for TIV). At 4 years of follow-up, analyses were performed on 1,231 participants for hypertension, 1,222 participants for HV (by also excluding outliers for TIV) and 951 participants for WMH (by also excluding outliers for TIV).

All individuals with HV available at baseline (n = 1,576) were considered in a sensitivity analysis to confirm HV results.

### Covariates

In the mediation analysis, age, gender and educational level (coded in 6 categories) were included as covariates. Total intracranial volume (TIV) was also included as covariate in mediation analyses of MRI metrics (i.e. WMH, HV, and HV atrophy rate).

### Statistical analysis

For the comparison of patient characteristics between demented and non-demented groups, categorical variables were compared using chi-square statistics. Age was compared using t-test. Comparisons of *APOE4* status, WMH volume, HV and HV atrophy rate between demented and non-demented groups were performed using logistic regression analyses adjusted for age, sex, education level and for TIV for MRI variables. Mediation analyses were performed to identify mechanisms underlying the association between *APOE4* and dementia, as shown in Figure 1. Path [a] refers to the associations between *APOE4* and potential mediators, as assessed by logistic or linear regressions depending on the studied mediator; Path [b] refers to associations between potential mediators and dementia; Path [c] refers to the total effect of *APOE4* on dementia risk, as assessed by a logistic regression. Path [c’] refers to the direct effect of *APOE4* on dementia risk, adjusted for the mediators and assessed by a logistic regression. All regressions conducted within the mediation analyses were adjusted for age, sex and education level and were also adjusted for TIV if the potential mediator was an MRI variable. Bootstrap method was used to estimate the mediated proportion of the association between *APOE4* and dementia.

**Figure 1.**
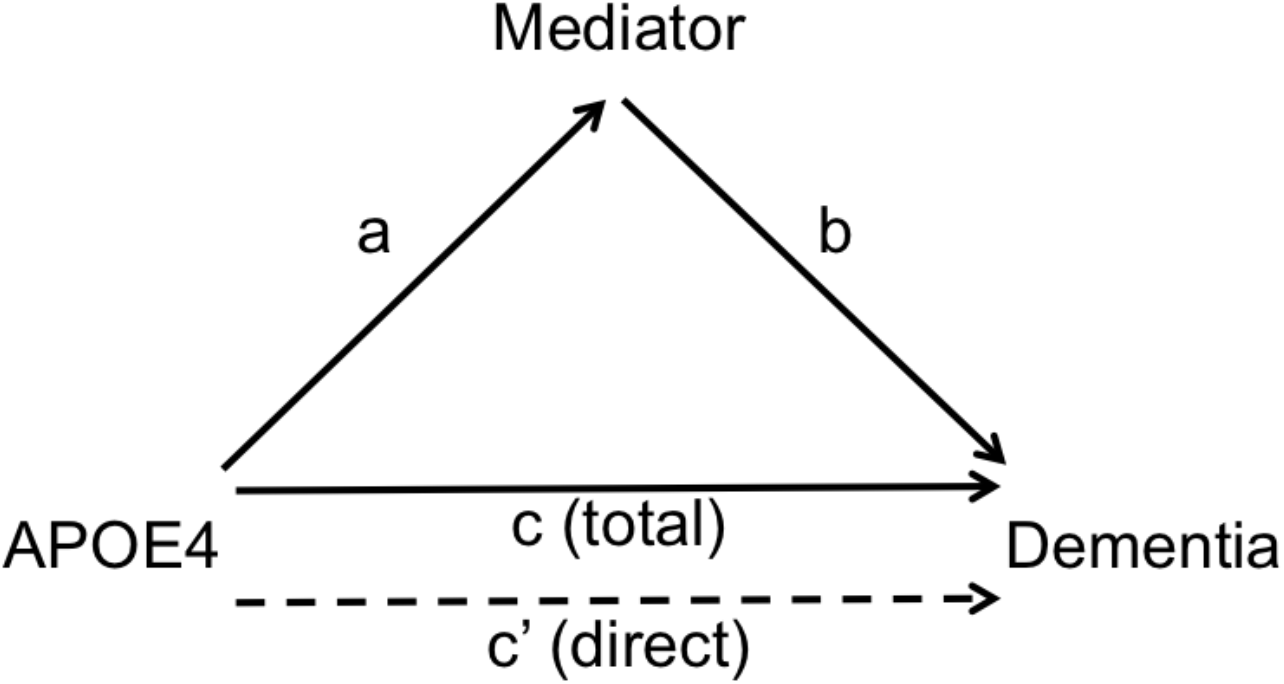
Mediation analysis of the association between *APOE4* and dementia. Path [a]: associations between *APOE4* and potential mediators; Path [b]: associations between potential mediators and dementia; Path [c]: total effect of *APOE4* on dementia risk; Path [c’] : direct effect of *APOE4* on dementia risk, adjusted for the mediator.

All statistical analyses were performed in R. The R package [mediation] was used for mediation analyses ^23^.

### Data availability statement

Data access can be requested through the 3C Scientific Committee (http://www.three-city-study.com).

## Results

### Participant characteristics

Participant characteristics at baseline are summarized in Table 1. Among the 1,240 participants at baseline, 76 (6.1%) developed a dementia during the 12 years of follow-up (11,069 persons-years, mean follow-up time = 8.9 years, standard deviation (SD) = 2.6 years, considering the last visit for non-demented participants and the first visit with a dementia diagnosis for demented participants). On average and as expected, demented participants were older at baseline than non-demented participants (74.3 vs 71.8 years, respectively, p < 0.0001) and had a lower education level (p = 0.005). As expected, demented participants had a lower HV compared to non-demented participants (p = 0.002), a higher HV atrophy rate (p < 0.0001), and a higher WMH volume (p = 0.03), and an association between *APOE4* and dementia was also observed (odds ratio (OR) = 1.94, 95% confidence interval (CI) = [1.19, 3.08], p=0.006). Demented and non-demented participants did not significantly differ with regard to gender, BMI, smoking status, diabetes, hypercholesterolemia, and hypertension.

**Table 1.** Participant characteristics at baseline. Abbreviations : SD : standard deviation ; BMI : body mass index ; HV : hippocampal volume ; WMH : white matter hyperintensities (1) p-values were adjusted for age, sex, education level and total intracranial volume, (2) p-values were adjusted for age, sex and education level.

| Characteristics | Demented | Non-demented | p-value |
| --- | --- | --- | --- |
| **Number of participants (n)** | 76 | 1164 |  |
| **Age (y), mean (SD)** | 74.3 (4.1) | 71.8 (3.9) | < 0.0001 |
| **Female, n (%)** | 54 (71.1) | 736 (63.2) | 0.21 |
| **Education, n (%)** |  |  |  |
| **1. No education** | 4 (5.3) | 11 (0.9) | 0.005 |
| **2. Elementary school** | 31 (40.8) | 357 (30.7) |  |
| **3. Professional certificate** | 8 (10.5) | 151 (13.0) |  |
| **4. Middle school** | 13 (17.1) | 216 (18.6) |  |
| **5. High school** | 5 (6.6) | 152 (13.1) |  |
| **6. College** | 15 (19.7) | 277 (23.8) |  |
| **Smoking, n (%)** |  |  |  |
| **Never smoker** | 54 (71.1) | 731 (62.8) | 0.26 |
| **Past smoker** | 20 (26.3) | 364 (31.3) |  |
| **Current smoker** | 2 (2.63) | 69 (5.9) |  |
| **BMI (kg/m2), mean (SD)** | 25.6 (3.5) | 25.0 (3.4) | 0.20 |
| **Diabetes, n (%)** | 7 (9.2) | 89 (7.7) | 0.79 |
| **Hypercholesterolemia, n (%)** | 43 (56.6) | 675 (58.0) | 0.9 |
| **Hypertension, n (%)** | 61 (80.3) | 874 (75.1) | 0.38 |
| **Total HV (cm3), mean (SD)** | 6.22 (0.79) | 6.67 (0.77) | 0.002<sup>(1)</sup> |
| **Total WMH volume (cm3), mean (SD)** | 5.21 (2.37) | 4.42 (2.02) | 0.03<sup>(1)</sup> |
| **HV Atrophy rate (cm3/year), mean (SD)** | -0.11 (0.06) | -0.06 (0.06) | < 0.0001 |
| **APOE4 (%)** | 16.4 | 10.9 | 0.006<sup>(2)</sup> |

Participant characteristics analyzed at 4 years led to similar results, except that the association with educational level did not reach significance (p=0.18). Demented participants were older than non-demented participants (77.8 vs 75.4 years, SD = 4.0 and 3.9 years, respectively, p < 0.0001) and had a lower HV (mean HV = 5.87 vs 6.46 cm3, SD = 0.76 and 0.83 cm3, respectively, p < 0.0001) and a higher WMH volume (mean WMH volume = 5.97 vs 5.04 cm3, SD = 2.6 and 2.38 cm3, respectively, p = 0.01). An association between *APOE4* and dementia was also observed (OR=2.07, 95% CI = [1.26-3.33], p=0.003).

### Mediation analyses

The results of the mediation analyses at baseline are summarized in Table 2. Among the 8 mediators tested, only HV was identified as a significant mediator of the association between *APOE4* and dementia and accounted for 6.7% of the observed association (p=0.03). Both association of *APOE4* with HV (β = -0.08, SD = 0.04, p=0.04) and association of HV with dementia (β = -0.65, SD = 0.21, p=0.002) were significant. The total effect of the association of *APOE4* with dementia was 0.67 (SD=0.24), whereas the direct effect, including HV as a covariate, was 0.62 (SD=0.25). *APOE4* was associated with hypercholesterolemia (p=0.004) but hypercholesteromia was not associated with dementia in our sample (p=0.58). No mediation analyses could be undertaken for the others factors due to the lack of their significant association with *APOE4* in our sample.

**Table 2.** Mediation effects of potential mediators, evaluated at baseline, of the association between *APOE4* and dementia. For the definition of the paths, see material and methods and figure 1. Abbreviations: SD: standard deviation ; HV : hippocampal volume ; WMH : white matter hyperintensities All regressions analyses were adjusted for age, sex and education level. TIV was also included a covariate for mediation analyses of MRI mediators. Smoking 1: current vs never smoker model; Smoking 2: current versus former/never smoker model; Smoking 3: current/former versus never smoker model.

| Potential mediators | Path a |  | Path b |  | Total effect | Direct effect | p-value | Mediation ratio (%) |
| --- | --- | --- | --- | --- | --- | --- | --- | --- |
|  |  |  |  |  | (Path c) | (Path c') |  |  |
| | $\beta \pm SD$ | p-value | $\beta \pm SD$ | p-value | $\beta \pm SD$ | $\beta \pm SD$ | | |
| *Lifestyle factors* |  |  |  |  |  |  |  |  |
| Smoking 1 | -0.14 ± 0.32 | 0.65 | - | - | - | - | - | - |
| Smoking 2 | 0.12 ± 0.16 | 0.46 | - | - | - | - | - | - |
| Smoking 3 | -0.32 ± 0.31 | 0.30 | - | - | - | - | - | - |
| BMI | -0.15 ± 0.22 | 0.48 | - | - | - | - | - | - |
| *Co-morbidities* |  |  |  |  |  |  |  |  |
| **Diabetes** | -0.25 ± 0.26 | 0.34 | - | - | - | - | - | - |
| **Hypercholesterolemia** | 0.40 ± 0.14 | **0.004** | -0.14 ± 0.25 | 0.58 | - | - | - | - |
| **Hypertension** | 0.16 ± 0.16 | 0.30 | - | - | - | - | - | - |
| *MRI* |  |  |  |  |  |  |  |  |
| **WMH volume** | -0.02 ± 0.04 | 0.61 | - | - | - | - | - | - |
| **HV** | -0.08 ± 0.04 | **0.04** | -0.65 ± 0.21 | **0.002** | 0.67 ± 0.25 | 0.62 ± 0.25 | **0.03** | 6.7% |
| **HV atrophy rate** | -0.006 ± 0.004 | 0.11 | - | - | - | - | - | - |

Analyses using mediators evaluated at 4 years of follow-up confirmed these results, with a mediated proportion of 12.4% for HV (p<0.0001). Similar to the baseline analyses, no mediation analyses could be undertaken for hypertension and WMH volume evaluated at 4 years due to their lack of significant association with *APOE4* in our sample (p = 0.65 and p = 0.75, respectively)

The HV results at baseline were also confirmed in a sensitivity analysis by including all individuals with an HV available at baseline and HV accounted for 5.8% of the observed association (p=0.03).

## Discussion

We report here a role for HV as a partial mediator of the association between *APOE4* and dementia, accounting for 6.7% of the association. Among all mediators tested, HV was the only variable meeting all criteria for a mediation effect on the well-established association of *APOE4* with dementia ^3^. In our population, *APOE4* was indeed significantly associated with HV and HV was significantly associated with dementia. Integrating HV as a covariate in the regression model testing the association of *APOE4* with dementia led to a partial decrease in the reported effect, as expected for a mediator. The mediation effect was even higher (12.4%) when considering only participants who developed dementia after 4 years of follow-up. This is probably due to the older age of these patients and reflects the continuous rapid decrease of HV in demented patients compared to the natural decrease observed in normal aging ^24^. At first sight, the mediation effect seems to be low considering the role of *APOE4* and HV in AD. However, it has been reported that *APOE4* is also associated with cerebral Aβ and Tau aggregation ^25, 26^. Those mechanisms could be more valid cerebral mediators than HV itself. It would then be interesting to study them as potential mediators of the association between *APOE4* and dementia.

APOE is a glycoprotein ^27^ playing a critical role in the transport of cholesterol and other lipids and is known to be involved in injury repair in the brain^28^. In the central nervous system, it is expressed in astrocytes, microglia and stressed neurons ^29 30^. Mice studies seemed to show a role of the main AD genetic risk factor, *APOE4* ^2^, in the hippocampus, mice carrying *APOE4* having an impaired hippocampal neurogenesis ^31^. Reports also showed that in the brain of AD individuals, *APOE4* is associated with increased synaptic accumulation of Aβ oligomers ^32^, a biomarker of AD pathology, and synaptic degeneration ^33^. Blocking the interaction between APOE and Aβ also reduced intra-neuronal accumulation of Aβ and inhibited synaptic degeneration ^34^.

In our population, hypercholesterolemia was also a potential mediator associated with *APOE4*. However, it was not significantly associated with dementia in our sample. It was reported that hypercholesterolemia might be a risk factor for dementia but at a younger age ^11^. Absence of significant results might thus be due to a late assessment of hypercholesterolemia, which should have been assessed rather at young adulthood or middle age, or could be due to a lack of statistical power.

None of the other tested potential mediators were associated with *APOE4* in our population and therefore, were not included in mediation analyses. In a study using the whole 3C population and dementia status assessed at 8 years of follow-up, no difference in smoking status and in the mean level of cholesterol were observed between individuals with an incident dementia and non-demented individuals. However, differences were observed regarding hypertension and diabetes status ^35^. The non-association observed in our study for those two potential mediators could therefore be a result of the small size of our population. Our study has thus some limitations. Applying strict inclusion criteria resulted in a sample of 1,240 out of the 1,434 individuals with MRI data, and among them, only 76 had developed a dementia after up to 12 years of follow-up. Moreover, some factors were relatively uncommon in our population, such as diabetes (96 individuals), thus limiting the statistical power of our analyses and explaining some of the negative results that we observed. These inclusion criteria also probably introduced some selection bias leading to a population less representative of the general population. Indeed, in the Three-City Study, individuals with MRI were younger, healthier and more educated compared to individuals without MRI ^20^.

Nevertheless this study has several strengths. Participants included in this study have been followed-up on many years, with a mean follow-up time close to 9 years. All patients with MRI data came from a unique center and all baseline and follow-up structural brain scans were acquired using the same MRI machine and the same acquisition protocol ^20^, avoiding MRI metrics bias due to inter-centers and inter-machines variability.

In conclusion, hippocampal volume, a marker of neurodegeneration, seems to be involved in the association of *APOE4* with dementia. Other studies are needed to confirm this result.

## Data Availability

Data access can be requested through the 3C Scientific Committee (http://www.three-city-study.com)

## Acknowledgements

The authors thank all participants from the Three-City Study. The 3C Study supports are listed on the Study website (www.three-city-study.com)

